# Expert Surgeons and Deep Learning Models Can Predict the Outcome of Surgical Hemorrhage from One Minute of Video

**DOI:** 10.1101/2022.01.22.22269640

**Authors:** Dhiraj J Pangal, Guillaume Kugener, Yichao Zhu, Aditya Sinha, Vyom Unadkat, David J Cote, Ben Strickland, Martin Rutkowski, Andrew Hung, Animashree Anandkumar, X.Y. Han, Vardan Papyan, Bozena Wrobel, Gabriel Zada, Daniel A Donoho

## Abstract

**Background:** Major vascular injury resulting in uncontrolled bleeding is a catastrophic and often fatal complication of minimally invasive surgery. At the outset of these events, surgeons do not know how much blood will be lost or whether they will successfully control the hemorrhage (achieve hemostasis). We evaluate the ability of a deep learning neural network (DNN) to predict hemostasis control ability using the first minute of surgical video and compare model performance with human experts viewing the same video.

**Methods:** The publicly available SOCAL dataset contains 147 videos of attending and resident surgeons managing hemorrhage in a validated, high-fidelity cadaveric simulator. Videos are labeled with outcome and blood loss (mL). The first minute of 20 videos was shown to four, blinded, fellowship trained skull-base neurosurgery instructors, and to SOCALNet (a DNN trained on SOCAL videos). SOCALNet architecture included a convolutional network (ResNet) identifying spatial features and a recurrent network identifying temporal features (LSTM). Experts independently assessed surgeon skill, predicted outcome and blood loss (mL). Outcome and blood loss predictions were compared with SOCALNet.

**Results:** Expert inter-rater reliability was 0.95. Experts correctly predicted 14/20 trials (Sensitivity: 82%, Specificity: 55%, Positive Predictive Value (PPV): 69%, Negative Predictive Value (NPV): 71%). SOCALNet correctly predicted 17/20 trials (Sensitivity 100%, Specificity 66%, PPV 79%, NPV 100%) and correctly identified all successful attempts.

Expert predictions of the highest and lowest skill surgeons and expert predictions reported with maximum confidence were more accurate. Experts systematically underestimated blood loss (mean error −131 mL, RMSE 350 mL, R^2^ 0.70) and fewer than half of expert predictions identified blood loss > 500mL (47.5%, 19/40). SOCALNet had superior performance (mean error −57 mL, RMSE 295mL, R^2^ 0.74) and detected most episodes of blood loss > 500mL (80%, 8/10).

In validation experiments, SOCALNet evaluation of a critical on-screen surgical maneuver and high/low-skill composite videos were concordant with expert evaluation.

**Conclusion:** Using only the first minute of video, experts and SOCALNet can predict outcome and blood loss during surgical hemorrhage. Experts systematically underestimated blood loss, and SOCALNet had no false negatives. DNNs can provide accurate, meaningful assessments of surgical video. We call for the creation of datasets of surgical adverse events for quality improvement research.

## Introduction

Major bleeding complications during minimal access, endoscopic or robotic-assisted surgery can impair visualization and requires immediate action to control.^1,2^ Despite maximal efforts, including the conversion from minimally invasive to ‘open’ surgery, 13-60% of major vascular injuries result in patient death.^2–6^ Surgeon assessments of the likelihood of achieving hemostasis and the need for blood transfusion should be made immediately; however, inexperience, inability ^7–11^and stress ^1,3,12,13^ impair decision-making, and surgeon self-assessments of the likelihood of controlling an unexpected vascular complication are uncorrelated with their actual performance.^14^ Inaccurate predictions of blood loss and task outcome risk patient harm by delaying changes in technique, aid from surgical colleagues, or transfusion of blood products. Rather than waiting for a patient’s clinical deterioration, early prediction of difficulty at achieving hemostasis and high-volume blood loss using computer vision (CV) techniques could optimize patient outcomes.

We created SOCAL (Simulated Outcomes following Carotid Artery Laceration), a video dataset of attending and resident surgeons (otorhinolaryngologists and neurosurgeons) controlling life-threatening internal carotid artery injury (ICAI) in a validated, high-fidelity bleeding cadaveric simulator.^14–18^ Carotid injury is a catastrophic complication of endonasal surgery and results in up to 30% mortality, similar to vascular injuries during minimally-invasive abdominal and thoracic surgery.^5,19,20^ In prior work, we applied artificial intelligence (AI) methods to SOCAL video and developed tools that quantify blood loss and measure surgeon performance metrics from video.^21,22^ Using these tools, we showed that video contains signals of surgical task outcome, but we do not know whether the model can detect predictive signals early in a bleeding episode, nor its performance compared to gold-standard human experts We provided human experts (fellowship trained skull-base neurosurgeons) with the first minute of 20 videos from SOCAL (‘Test Set’) and collected predictions of blood loss and task success over the entire unseen task. Experts’ predictions of outcome and blood loss established a benchmark of human performance. We then built a deep learning neural network (DNN) trained on the SOCAL video dataset (excluding the Test Set), called SOCALNet, and compared model performance on the Test Set to expert benchmarks. We validated SOCALNet predictions in subsequent experiments. To the authors knowledge this is the first comparison of DNN-derived surgical video outcome prediction to human experts viewing the same video.

## Methods

### Experimental Design

Experimental setup, data collection, consent and implementation parameters for the dataset are found in Appendix 1. Seventy-five surgeons ranging from junior trainees to world experts on endoscopic endonasal approaches (EEA) were recorded in a nationwide, validated, high-fidelity training exercise. Surgeons attempted to control an ICAI in a cadaveric head perfused with blood substitute. Performance data and intraoperative video was used to develop the SOCAL database.^14–18,23^ The SOCAL database was developed in concordance with previously published methods, and is publicly available.^23–25^ The SQUIRE reporting guidelines were followed.^26^ The study was approved by the IRB of the University of Southern California. All research was performed in accordance with relevant regulations/guidelines. No patient data was utilized therefore patient-level informed consent was waived. Participating surgeons’ consent was obtained for intraoperative video recording. Surgeon-expert consent was obtained.

### Datasets

The 147 videos in SOCAL were divided into a training set of 127 videos and a separate test set of 20 videos. Ten videos depicting successes and 10 of failure were initially chosen at random for the test set; ultimately, 11 success videos (and 9 failures) were used due to ease of video formatting. Videos were truncated after 60 seconds. Only videos in the test set were shown to experts for grading.

### SOCALNet Model Development

See eSupp1 for model code. Video was sampled at 1 frame-per-second (fps) and input into two layers, a feature generating layer and a temporal analysis algorithm (**Figure 1**). The output of the model was a binary prediction of surgical ability (trial success or failure) and estimated blood loss over the entire trial (in milliliters).

**Figure 1.**
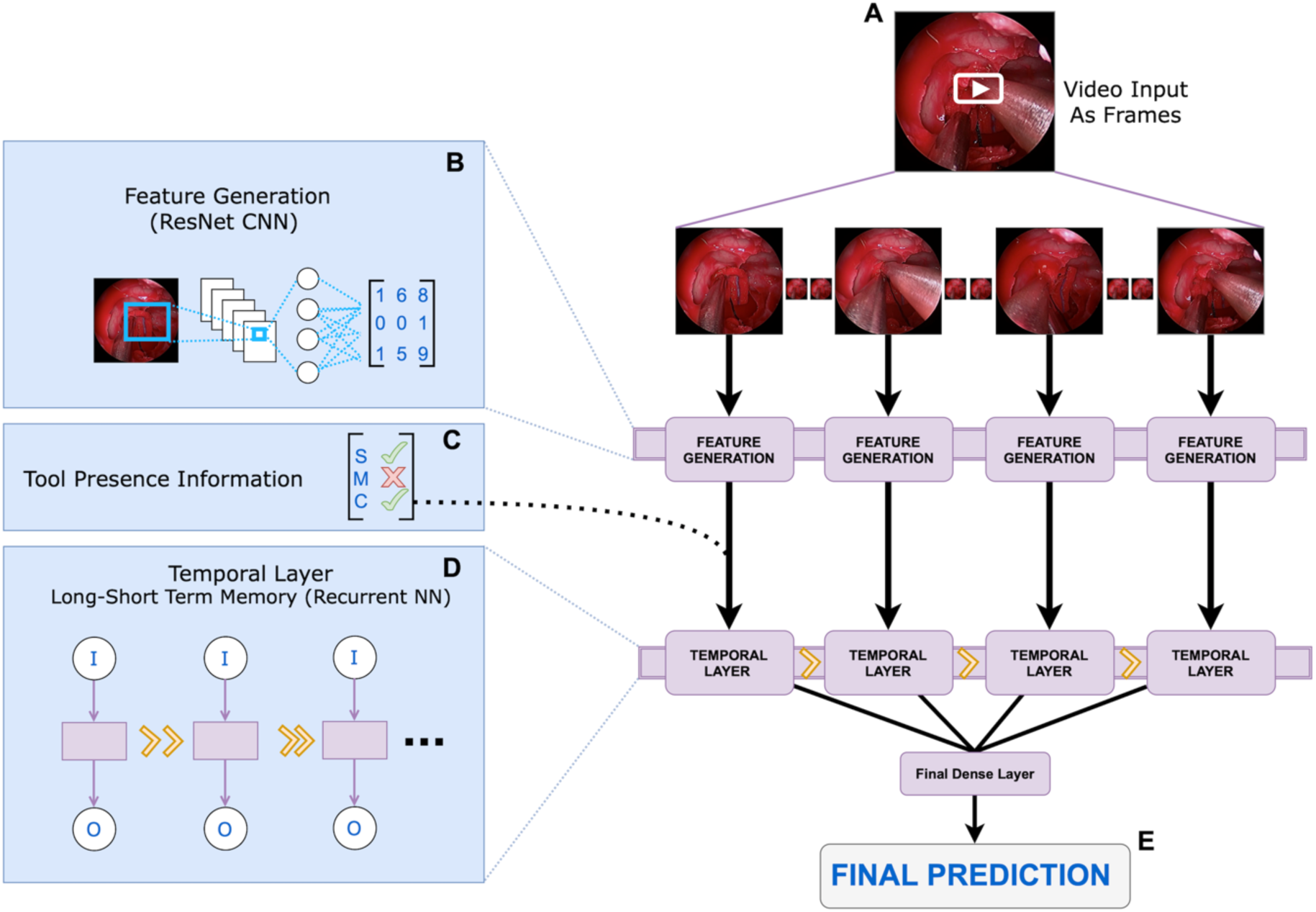
SOCALNet Architecture. Deep learning model used to predict blood loss and task success in critical hemorrhage control task. A) Video is snapshotted into individual frames. B) A pretrained ResNet convolutional neural network (CNN) is fine-tuned on SOCAL images from (A), to find features predictive of blood loss and task success in each individual frame. Output matrix from (B) and tool presence information (C) [e.g. Is suction (S) present? Yes (check); is Muscle (M) present? No (X), etc] is input into a temporal layer. D) Temporal layer: Long-short-term memory (LSTM) modified recurrent neural network allowing for temporal analysis across all frames. All LSTM predictions are consolidated in one dense layer and E) a final prediction of success/failure, and blood loss (in mL) is output

For the feature generator, we utilized a Residual Learning Neural Network (ResNet) model pretrained on the ImageNet 2012 classification dataset.^27,28^ ResNet is a single-stage convolutional neural network (CNN) which uses skip connections to allow for large networks with many layers to skip layers that hurt overall performance. ResNet has become ubiquitous for object detection and classification in computer vision (CV).^28^ The final three layers of the ResNet were retrained on SOCAL images to detect features indicative of blood loss or task success. Features from the penultimate layer of the ResNet and manual instrument annotations were passed into a bi-layer Long Short-Term Memory (LSTM) recurrent neural network.^29^ LSTM cells contain an input, output and forget gate, allowing the network to regulate the flow of information across cells. Instrument annotations alone are inadequate for outcome prediction; successful detectors incorporate instrument data and image features.^21^

### Expert Assessment

Experts were four skull base fellowship-trained neurosurgeon instructors in ICAI management. Experts watched the 20, one-minute test videos and provided: blood loss estimates (in mL), outcome predictions (success/failure), and surgeon grades (1-5 Likert scale, 1 represents novice and 5 represents master). Experts also reported self-confidence in their outcome prediction (1-5 Likert scale; 5 represents most confident). Prior to grading, experts watched anchoring videos of novice, average, and master performances with respective outcomes data. Anchoring videos were not contained in the Test-Set, and were chosen as representative videos of each skill level by adjudication by the study team. Grading sessions were conducted in double-blinded fashion by the lead author (DJP) and individual experts (BS, MR, GZ, DAD, referred to as S1-S4). Given high concordance, mean and mode are reported for experts (‘S’).

### Validation Analysis

We conducted two experiments to evaluate model and expert concordance. In experiment one, two videos were identified in the Test-Set which where a critical error occurred shortly after the 1-minute video sample concluded (i.e., not shown to the model or surgeons). The model and all surgeons predicted, incorrectly, that both videos were successes. A new, one minute clip was generated showing the critical error and its aftermath. These new clips were evaluated by one of the human experts and SOCALNet.

In a second experiment, the three best (least blood loss, successes) and worst (most blood loss, failures) videos were identified from within the Test-Set. Composite ‘best’ and ‘worst’ videos were constructed by combining the first 20 seconds of each of the three best and worst trials in each possible order permutation (6 ‘best’, 6 ‘worst’ videos). The twelve composite videos were then presented to SOCALNet.

### Statistical Analysis

Blood loss prediction was reported using mean error, root mean square error (RMSE), and Pearson’s correlation coefficients. Categorical inter-rater reliability was calculated using Cohen’s Kappa and Krippendorff’s alpha for more than two raters. Continuous inter-rater reliability was calculated using Pearson’s correlation coefficient and an inter-rater correlation coefficient (ICC) (>2 groups; using a two-way random effects ICC model).^30^ We used Fisher’s exact test for categorical comparisons. We performed analysis in Python with SciPy.^31^

## Results

**Table 1** lists predictions and ground truth data. There were 11 successful trials and 9 failed trials in the Test Set, with mean blood loss of 568mL (range 20-1640 mL, mean success=323 mL, mean failure=868 mL). Experts correctly predicted outcome in 55/80 predictions (69%, Sensitivity: 79%, Specificity: 56%). Expert predictions were concordant, with one dissent in 80 ratings (Fleiss’ kappa = 0.95). The average root mean square error (RMSE) for blood loss prediction of surgeons was 351 mL (mean error=-131mL, average R^2^ = .70). Expert ICC was high at 0.72.

**Table 1.** Results comparing Deep Learning Model with Expert Surgeons. SN: Sensitivity, SP: Specificity, M-S: Model-Surgeon. *: Kappa coefficient; **†**:inter-class coefficient; ‡: Inter-Surgeon Agreement: Success/Failure= 0.95, Blood-Loss: 0.72

|  | Accuracy<br>(SN %, SP %) | RMSE<br>(R<sup>2</sup>) | M-S Agreement:<sup>*</sup><br>*Success/Failure* | M-S Agreement:<sup>†</sup><br>*Blood Loss* |
| --- | --- | --- | --- | --- |
| Ground Truth | 11 Success<br>9 Failures | - | - | Avg Blood Loss: 568<br>(Range:20-1640 ) |
| Model | 17/20 (85%)<br>(100, 66) | 295<br>(.74) | - | - |
| Expert Cohort | **55/80 (68.75)**<br>**(79, 56)** | **351**<br>**(.70)** | **.43<sup>‡</sup>** | **0.73<sup>‡</sup>** |
| Surgeon 1 | 13/20 (65%)<br>(73, 55) | 306<br>(.73) | .34 | .74 |
| Surgeon 2 | 14/20 (65%)<br>(81, 55) | 335<br>(.66) | .43 | .66 |
| Surgeon 3 | 14/20 (65%)<br>(81, 55) | 423<br>(.65) | .43 | .65 |
| Surgeon 4 | 14/20 (65%)<br>(81, 55) | 329<br>(.74) | .43 | .72 |

**Figure 2, and Supplemental Table 1** demonstrates the relationship between prediction confidence, surgeon skill and prediction accuracy. Experts were most accurate when maximally confident (5/5 confidence, accuracy 88%) or viewing a surgeon they rated as having minimal (Likert scale 1, accuracy 92%) or maximal skill (Likert scale 5, accuracy 79%). Predictions with non-maximal confidence (levels 2-4,) were only marginally better than chance (53%, p=0.02 compared to maximal confidence). Predictions of intermediate skill surgeons were also less accurate (levels 2-4, 63%, p=0.04 compared to composite 1/5 and 5/5 skill).

**Figure 2.**
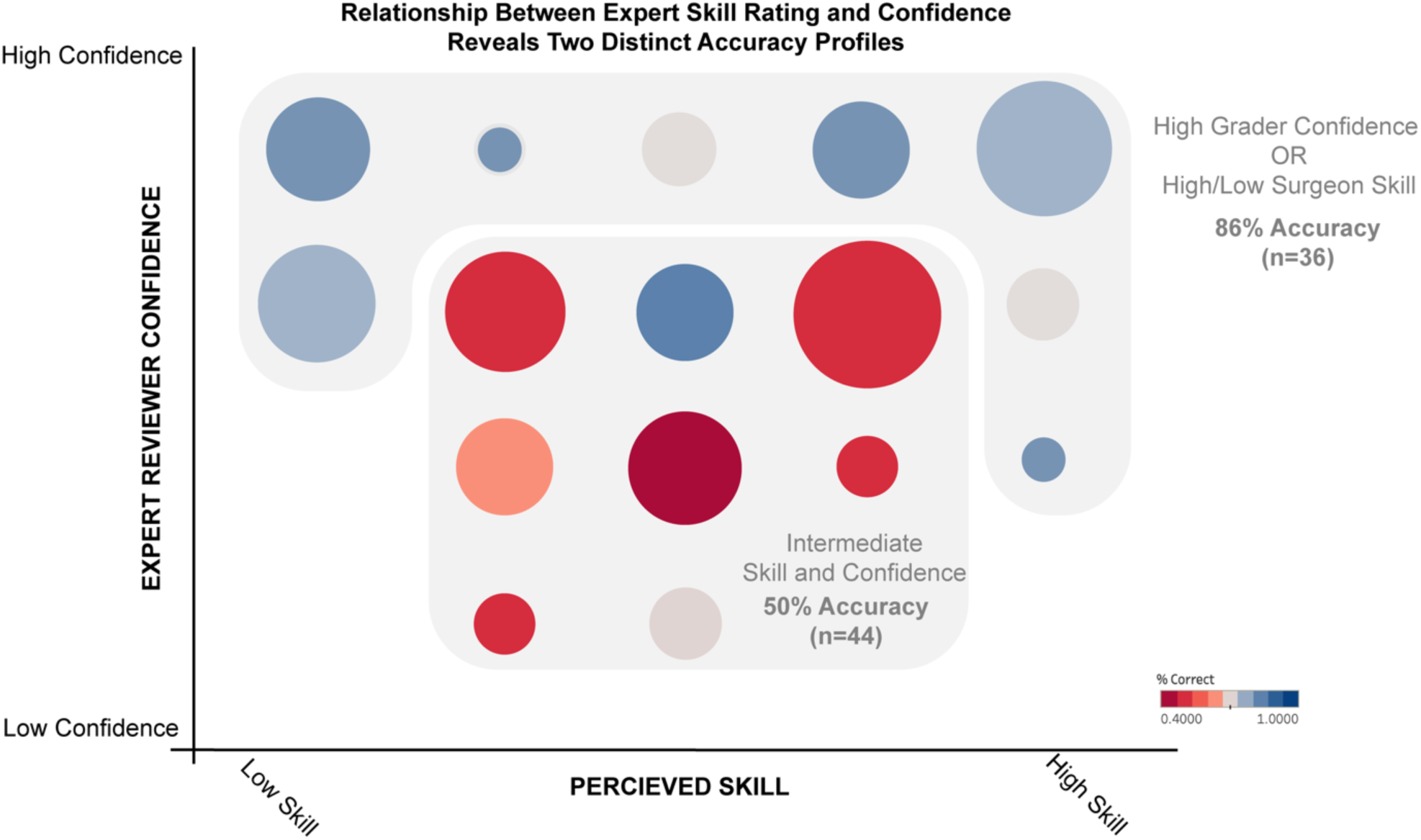
Association between expert confidence, surgeon skill level and accuracy of prediction. Experts are most accurate when viewing trials of surgeons with low or high skill, or where they (experts) are maximally confident. For those with moderate skill or when experts have moderate confidence, prediction accuracy is lower. Size of circle denotes number of trials. Color denotes accuracy.

SOCALNet correctly predicted outcome in 17/20 trials (85%, Sensitivity: 100%, Specificity: 66%), noninferior to surgeons (p=0.12). The model predicted blood loss with a RMSE of 295 mL (mean error=-57mL, R^2^=.74) (**Figure 3)**. The model and experts all predicted outcome correctly in 13/20 trials. In four trials, the model was correct and all experts incorrect, in one trial the model was incorrect, and all experts correct, and two trials all were incorrect (**Figure 4**). Correlation (R^2^) between blood loss estimates for the model, experts and ground truth are shown in **Supplemental Figure 1**, and range from 0.53-0.93. Correlation between the model and the average surgeon blood loss estimate was 0.73, ranging from 0.53 to 0.74 for individual surgeons (**Table 1**).

**Figure 3.**
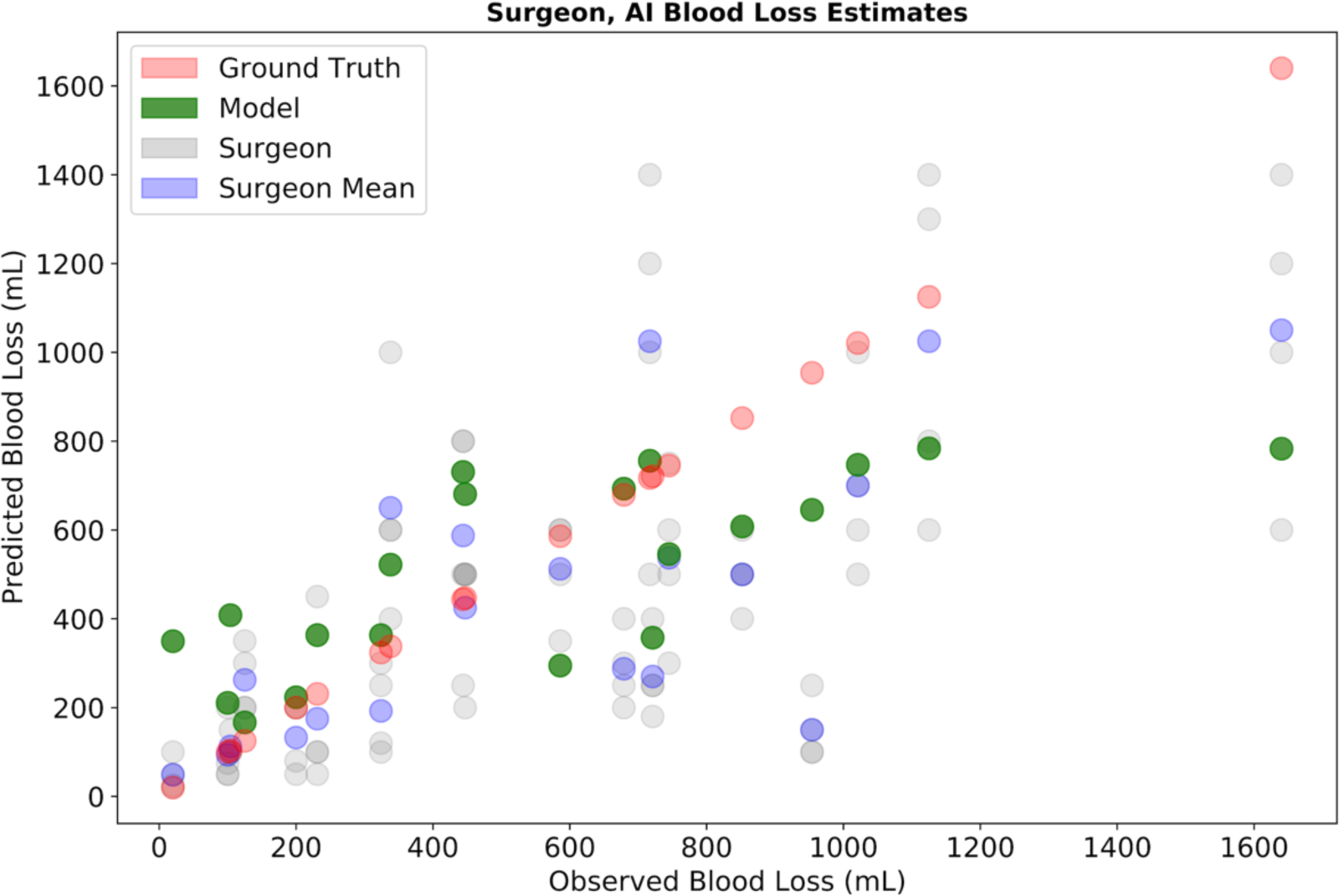
Expert and SOCALNet Blood Loss Quantification. Predicted versus observed blood loss estimations by individual surgeons (grey), surgeon mean (blue), and model (green). Red points represent measured blood loss (ground truth).

**Figure 4.**
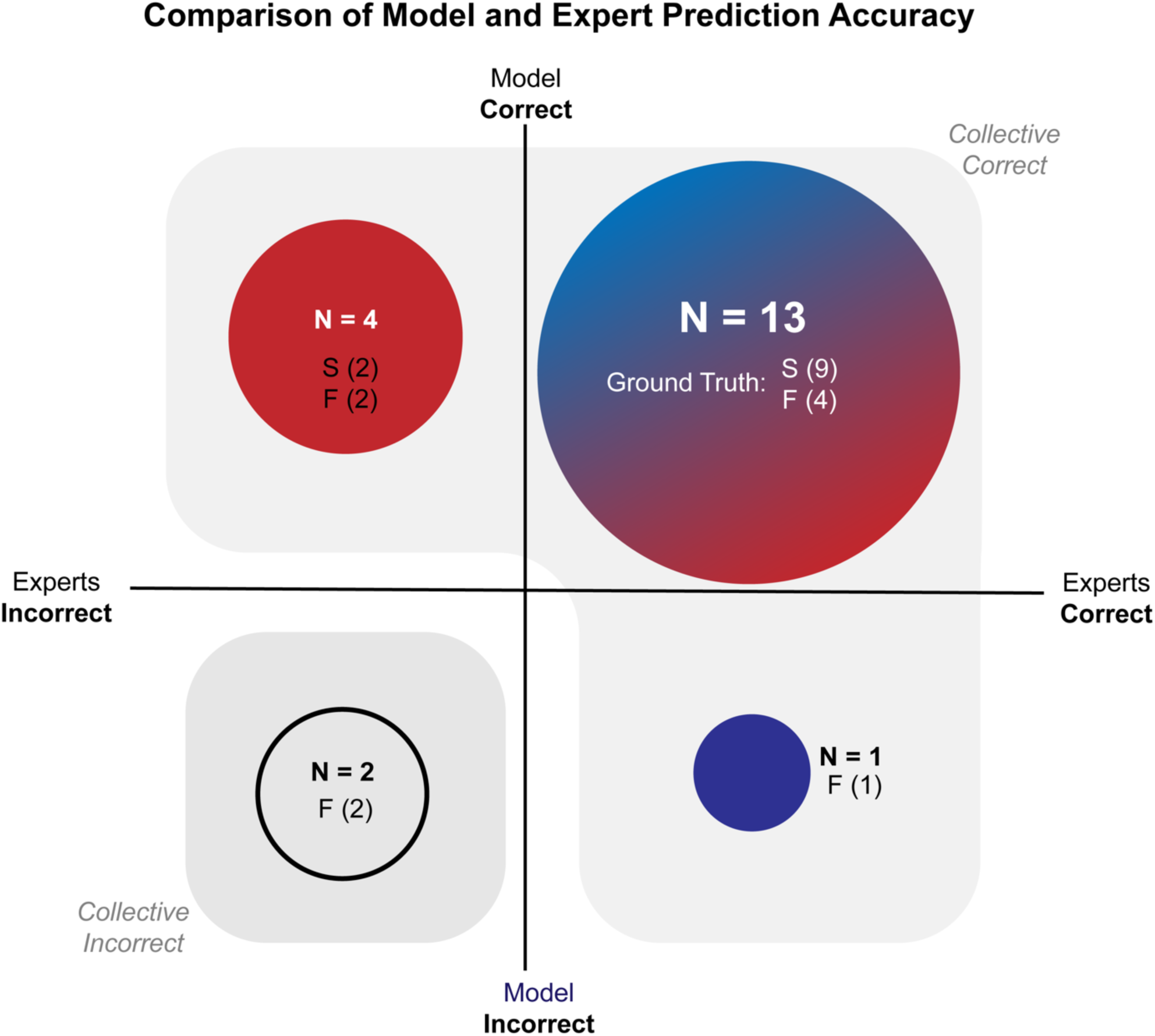
Outcome Predictions of Experts and SOCALNet. Outcomes of experts (Blue) and model (Red) in predicting task success using one minute of video. Circle size denotes number of trials (N). Success (S) and failure (F) denoted underneath each N. When the union of successful predictions is taken, the model+expert grouping would successfully predict outcome in 18/20 cases. In the 2 remaining cases (bottom left quadrant), a critical error took place following the cessation of the video and was evaluated in subsequent counterfactual experiments.

We then evaluated trials above the 50^th^ percentile for blood loss, where blood loss exceeded 500mL and transfusion might be needed. The model predicted a blood loss estimate above 500 mL in 80% (8/10) compared to experts 47.5% (19/40); this difference was not statistically significant (p=0.09).

## Exploratory Model-Validation

**Supplemental Table 2** reports model-validation experiments. In two trials, experts and SOCALNet predicted success, but the surgeon failed due to a critical error shortly after the end of the one-minute clip (therefore unseen by experts and SOCALNet). When we included the critical error, the model accurately predicted ‘failure’, as did an expert. In a second experiment, SOCALNet viewed six composite ‘Best’ trials and uniformly predicted success with low blood loss (328-473 mL); conversely, in six composite ‘Worst’ videos the model uniformly predicted failure with high blood loss (792-794mL).

## Discussion

To address the need for datasets depicting surgical adverse events we created SOCAL, a public video dataset of 147 attempts to control carotid injury in high-fidelity perfused cadavers. In this work we compared human expert predictions of outcome using one minute of video from 20 trials in the dataset to those of a DNN (SOCALNet). Compared to expert benchmarks, SOCALNet met or surpassed expert prediction performance, despite its relatively primitive architecture and small training data size relative to CV tasks. We synthesized counterfactual videos of excellent and poor surgeon performance to challenge SOCALNet, and it correctly predicted the outcomes in these challenges. SOCALNet and other CV methods can aid surgeons by quantifying and predicting outcome during surgical events, and in automatic video review. The absence of video datasets containing adverse events is a critical unmet need preventing the development of predictive models to improve surgical care.

### Benchmark Performance of Human Experts

Expert predictions were highly concordant, indicating that experts detected similar signals of blood loss and outcome (cross-correlation: R^2^ = 0.74 −0.93, Kappa for success prediction=0.95). Experts had uniform definitions of success (hemostasis) and were familiar with the stepwise progression of a well-described technique.^18,32^ Thus, it is reasonable to conclude that using the first minute of video of a bleeding event, human experts detect signals predictive of blood loss and task outcome.

Although experts had reasonably accurate outcome and blood loss predictions (69% accuracy, R^2^ =0.7), experts systematically overestimate surgeon success and underestimate bleeding: 4/6 of expert errors were false ‘success’ predictions, experts systematically underestimated blood loss by 131 mL and experts failed to identify 52% of high blood loss (above 500 mL) events. This post-hoc cutoff of 500mL represents a potential clinical marker of need for transfusion. The tendency for human experts to underestimate blood loss is well documented,^33–36^ corroborated by our findings, and may result in delayed recognition of life-threatening hemorrhage.

To validate individual ratings, we asked experts to provide their confidence in each prediction, and perceived skill rating of the participating surgeon. Maximally confident predictions were more likely to be correct, as expected from prior work.^33,34,37^ Similarly, predictions were most accurate when evaluating highest and lowest-skilled surgeons (skill rating 1 or 5), but scarcely better than chance when evaluating intermediate surgeons. Intermediate skill surgeons comprised half of all surgeons and may benefit greatly from performance assessments.

During a real vascular injury, estimation ability of the average surgeon is likely to be inferior to our experts calmly rating a single stereotyped task after training with videos of known blood loss. Experts’ systematic underestimation of blood loss and struggle to assess performance of intermediate surgeons represents a chasm in surgeon-assessment proficiency. Surgical patients may benefit from novel methods that improve on these benchmarks.

### SOCALNet Performance Compared to Experts

We designed a primitive deep-learning architecture containing a standard CNN and a recurrent neural network, which we call SOCALNet. We provided SOCALNet with short videos from a much smaller training dataset than is customary in CV. Despite these disadvantages, SOCALNet made statistically non-inferior (and numerically superior) outcome predictions and superior blood loss predictions compared to human experts. SOCALNet’s predictions of blood loss had a smaller mean underestimation and standard error. Unlike experts, SOCALNet predictions were accurate for intermediate-skill surgeons.

The advantages of SOCALNet support the development of computer vision tools for surgical video review and as potential teammates for surgeons.^38^ SOCALNet demonstrates that CV models can provide accurate, clinically meaningful analyses of surgical outcome from video. Future models could leverage the vast but largely untapped collections of surgical videos. Workflows developed in building SOCALNet can guide model deployment for other surgical adverse events. Human-AI teaming is a validated concept in other domains.^39–41^ A SOCALNet-and-expert combined team (with model as a tiebreaker, particularly when expert confidence was low) would have generated 18/20 correct predictions. Furthermore, the only two inaccurate predictions from this teaming occurred when a critical error was made after the video ceased, and these errors were detected by the model and experts. If utilized at scale, AI-driven video analysis may quantify comparisons of surgical technique, provide real-time feedback for trainees, or provide guidance during rare scenarios a surgeon may not have encountered (e.g. vascular injury) but the model has been trained on.^38^

SOCALNet has room for improvement. For adverse events, the 1) accurate estimation of high-volume blood-loss and 2) detection of task failures may be prioritized as exsanguination is life-threatening. SOCALNet blood loss predictions exhibited more robust central tendency than experts, resulting in better predictions for typical performances. However, when grading edge cases of the two worst surgeons in the Test Set, SOCALNet underestimated blood loss (absolute error of 790-800 mL on videos exceeding 1.5L of blood loss). In predicting failure (specificity), both experts and SOCALNet showed limitations (Specificity= 0.56, 0.66 respectively); however, improving expert predictions are challenging, and most surgeons are non-experts. Accordingly, applying CV optimization techniques to AI models (e.g. cost-sensitive classification, oversampling) may be preferred.^42,43^

### Surgical Adverse Event Video Datasets: An Unmet Need in Surgical Safety

A growing body of evidence supports the quantitative analysis of surgical video.^22,44–47^ One fundamental discovery has been the detection of signals in surgical video that predict patient outcome: surgeons have heterogeneous skill resulting in heterogeneous outcomes.^14,44,45,48^ Although low-skill surgeons are more likely to have adverse intraoperative events, video of these events has not been systematically studied. Instead of studying surgical video, studies describe adverse events using textual medical records, radiography, and laboratory results. Analysis of these extra-operative records and correlations with pre-operative risk factors and post-operative management can be useful.^49–53^ However, this research omits a crucial determinant of the outcome of the surgical patient: the surgical event itself. This omission limits root-cause analysis to only the extra-operative universe and prevents evaluation of the technical maneuvers and patient anatomic conditions that make adverse events more likely. Unlike textual records, surgical video depicts all visualized surgeon movements and patient anatomy, making video uniquely suited for the study of operative events. The results of the present study begin to demonstrate the value of studying video of surgical adverse events.

We propose the creation of large, multi-center datasets of surgical videos that includes adverse events.^54,55^ Video datasets of surgical adverse events can be leveraged using predictive models (e.g., SOCALNet) which can detect intraoperative events, evaluate performance and quantify technique. This study was supported the North American Skull Base Society, whose mission is to promote scientific advancement, share outcomes data for education and to advance outcomes research. Groups such as the Michigan Bariatric Surgery Collaborative and the Michigan Urologic Surgery Improvement Consortium have conducted similar work and we hope to call their attention to adverse events in addition to routine procedures.^56,57^ National organizations capable of soliciting large bodies of data should prioritize collecting adverse event videos and apply technical innovations adopted by other medical fields to ensure privacy and confidentiality.^58–60^ National organizations can also facilitate the scaling of expert labeling. Small groups face long delays in accruing sufficient cases and labeling video. In this study, despite a long term track record of collaboration amongst our team, it required two months for our experts to review 20 minutes of aggregated video.^61^ Collaborative efforts may be able to require video review as a condition of membership.

Finally, high-fidelity simulation enables analysis of rare surgical events. Curating 150 videos of real carotid injuries would require tens of thousands of cases, an impossible task without streamlined data-sharing mechanisms; using perfused cadavers and real instruments we collected hundreds of observations of this otherwise rare event. Videos in the simulated environment can complement surgical video datasets that otherwise depict thousands of uncomplicated cases and only a few rare events.^14,15,17,18,62–65^ As more surgical video datasets are developed, we can follow the ‘sim-to-real’ process where models are trained on virtual data and then fine-tuned and validated in the real environment.^66–68^

## Limitations

Our study has several limitations. First, validation on clinical video is a clear next step, although accruing a corpus of carotid injury video would likely require substantial national efforts. Second, results from carotid injuries may not transfer to other vascular injuries, and vascular injuries differ from other adverse events. Rather than diminishing our results, these complementary challenges showcase the depth of unmet need within surgical-video data science. Separately from these study design limitations, SOCALNet ingests ground truth tool annotations as input, which requires pre-processing of data and is thus not fully automated.^69–71^ The lack of curated surgical video datasets remain a major limitation for future work.

## Conclusion

Experts and a neural network can predict the outcome of surgical hemorrhage from the first minute of video of the adverse event. Neural network-based architectures can already achieve human or supra-human performance at predicting clinically relevant outcomes from video. To improve outcomes of surgical patients, advances in quantitative and predictive methods should be applied to newly collected video datasets containing adverse events.

## Supporting information

SOCAL Appendix

## Data Availability

All data produced are either available online on figshare, or upon reasonable request to the authors.

https://doi.org/10.6084/m9.figshare.15132468.v1

## Data Availability

The datasets generated during and/or analyzed during the current study are available in the *figshare* repository, link: https://doi.org/10.6084/m9.figshare.15132468.v1

---

**Supplemental Figure 1.**
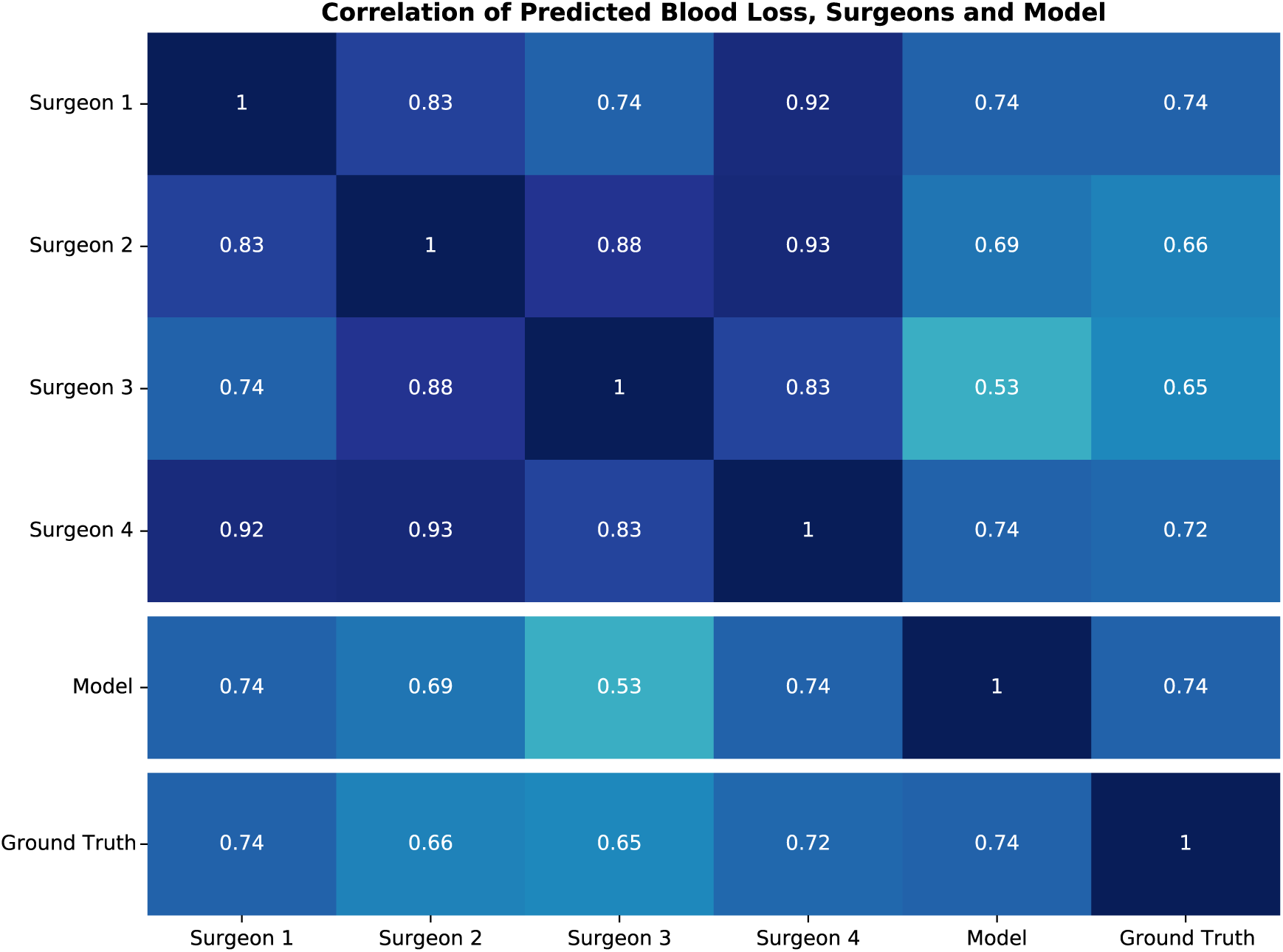
Correlation (R^2^) between blood loss prediction from all 4 expert surgeon graders, model, and ground truth data.

