## Supplemental Tables for "Expert Surgeons and Deep Learning Models Can Predict the Outcome of Surgical Hemorrhage from One Minute of Video"

**Supplemental Table 1**

| **Skill Rating** | **Number of**  **Ratings** | **Prediction Accuracy** | **Mean Absolute Error (Blood, mL)** |
| --- | --- | --- | --- |
| 1 | 13 | 0.92 | 461 |
| 2 | 16 | 0.56 | 195 |
| 3 | 18 | 0.61 | 236 |
| 4 | 19 | 0.63 | 297 |
| 5 | 14 | 0.79 | 131 |
| *Significance** |  | 0.21  0.04 (Pooled) | 0.002 |
| **Expert Confidence** |  |  |  |
| 1 | 0 |  |  |
| 2.0 | 5 | 0.60 | 269 |
| 3.0 | 15 | 0.53 | 179 |
| 4.0 | 35 | 0.63 | 337 |
| 5.0 | 25 | 0.88 | 200 |
| *Significance** |  | 0.08  0.02 (Pooled) | 0.07 |

Supplemental Table 1. Association between rated skill level (Likert scale, 1 = novice, 5 = master surgeon) and confidence (Likert scale, 1 = very uncertain, 5 = very confident) on accuracy of hemorrhage control prediction and blood loss estimates. *Significance testing: Chi-squared test was used for accuracy prediction (categorical), analysis of variance (ANOVA) used for continuous variable. Pooled calculations were conducted by aggregating ‘moderate’ ratings (2-4) and comparing to ‘extreme’ ratings (1 or 5). Model performance on ‘moderate’ trials: 80% (skill rating), 90% (confidence) (p=0.41, 0.14 respectively).

| **Cross-Stitch Trials** | **Ground Truth* (mL)** | **Blood Loss Prediction (mL)** | **Success Prediction** |
| --- | --- | --- | --- |
| ‘Best’ Version 1 | 75 | 462 | Success |
| ‘Best’ Version 2 | 75 | 417 | Success |
| ‘Best’ Version 3 | 75 | 328 | Success |
| ‘Best’ Version 4 | 75 | 364 | Success |
| ‘Best’ Version 5 | 75 | 386 | Success |
| ‘Best’ Version 6 | 75 | 473 | Success |
| ‘Worst’ Version 1 | 1262 | 793 | Failure |
| ‘Worst’ Version 2 | 1262 | 792 | Failure |
| ‘Worst’ Version 3 | 1262 | 794 | Failure |
| ‘Worst’ Version 4 | 1262 | 793 | Failure |
| ‘Worst’ Version 5 | 1262 | 792 | Failure |
| ‘Worst’ Version 6 | 1262 | 792 | Failure |
| **Shifted-Input Trials** | **Critical Error** | **Time Frame (s)** | **Prediction** |
| Video 1 | ~65 seconds | 0 – 60 | Success |
| Video 1 |  | 50 – 110 | Failure |
| Video 2 | ~65 seconds | 0 – 60 | Success |
| Video 2 |  | 50 - 110 | Failure |

Supplemental Table 2. Follow-up experiments conducted to investigate methodology of model. Cross-stitch trials had 20 second segments from the three best trials by blood loss stitched together to form a 1 min segment in various orders (‘Best’ Versions 1-6), and 20 seconds from the three worst trials by blood loss (‘Worst’ Versions 1-6). These would represent clear successes and clear failures and validates claims regarding what the model may be using to make predictions. Shifted input trials represent trials where surgeons and models were not shown the critical error, and all incorrectly assessed the surgeon. When given the critical error, the surgeons would clearly identify the trial as a failure, and we demonstrate similar performance in the model. *Ground truth data was derived from taking the average actual blood loss from the respective trials.
