## Supplementary material for "Expert Surgeons and Deep Learning Models Can Predict the Outcome of Surgical Hemorrhage from One Minute of Video": SOCAL Appendix

**Appendix A**

**SOCAL Dataset Methodology:**

Portions of the following sections are adapted from Donoho et al., 2021^1^, Kugener et al., 2021^2^. Data is publicly available at the following [link](https://doi.org/10.6084/m9.figshare.15132468.v1).

**Experimental Setup:**

Resident, fellow, and attending surgeons, including neurosurgery, and otorhinolaryngology–head and neck, were recruited for participation at nationwide educational courses between 2017 and 2020 (the North American Skull Base Society Annual Meeting, North American Skull Base Society Summer Skull Base Surgery Course, University of Southern California Annual Hands-On Comprehensive Neuro-Endoscopy Course, Emory Cranial Base Surgery Course, and Stryker Med-Ed Skull Cranial Surgery Course). The study was approved by the IRB of the University of Southern California.

A high-fidelity simulated operating room was constructed, including a surgical technician, surgical field, and simulated patient vital signs. A lightly embalmed human cadaveric head was prepared, and a standard endonasal endoscopic approach to the sella turcica was performed by study staff. Following cadaver perfusion at a standardized flow rate and physiological blood pressure using an artificial blood substitute, injury of the cavernous segment of the internal carotid artery (ICA) was induced by laceration. Participants were given standardized verbal instructions on the parameters, instruments, and goals of the simulation, but were not initially given specific technical instructions.

The protocol consisted of trial 1 (T1), followed by an educational intervention, and then trial 2 (T2) was performed. During T1 and T2, participants attempted to control the perfused ICAI using a variety of standard instruments and techniques (suction, cottonoid patties, and, ultimately, muscle patch control). Monitors showed simulated vital sign decompensation, and each trial ended when either hemostatic control was obtained using a muscle patch or simulated mortality occurred at 300 seconds (defined as ‘trial failure’). Time to hemostasis (TTH, in seconds) and blood loss (BL, in mL) were evaluated for each trial. After T1, subjects received specific feedback from one of the course instructors (endoscopic endonasal approach experts) and watched a standardized video of a senior author (G.Z.) explaining the recommended stepwise technique of ICAI management (Video 1). T2 was then performed with feedback.

**Data and Videos**

Intraoperative video was taken from the Karl Storz Video Neuro-Endoscope

used during each of these trials. A total of 143 videos from this nationwide educational intervention were recorded and saved. Videos were recorded at a frame rate of 30 frames per second (fps) and at a resolution of 1280x720 or 1920x1080. These videos are taken from multiple cadaveric heads, with different lighting, anatomy, laceration sites, camera resolutions, and brands of endoscopic instruments. The duration of the trials varies from 46 seconds to 5 minutes. These videos were down sampled from 30 frames-per-second (fps) to 1fps using ffmpeg, and were manually annotated to outline surgical instruments in each video frame using bounding boxes following previously published protocols using the open-sourced image annotation software VoTT.^3,4^

In conjunction with trial video recordings, “outcomes data” (e.g. blood loss, task success) and demographic data (e.g. training status, confidence) was recorded for each participant.

**This collection of annotated videos and corresponding surgeon demographics and performance data is termed the Simulated Outcomes Following Carotid Artery Laceration (SOCAL) Video Dataset^2^.**
